# A gene expression platform to predict benefit from adjuvant external beam radiation in resected non-small cell lung cancer

**DOI:** 10.1101/2021.01.31.21250879

**Authors:** Kamran A. Ahmed, Ben C. Creelan, Jeffrey Peacock, Eric A Mellon, Youngchul Kim, G. Daniel Grass, Bradford A Perez, Stephen A. Rosenberg, Thomas J. Dilling, Steven A. Eschrich, Alberto A. Chiappori, Javier F. Torres-Roca

## Abstract

**Background:** We hypothesized that the radiosensitivity index (RSI), would classify non-small cell lung cancer (NSCLC) patients into radioresistant (RR) or radiosensitive (RS).

**Methods:** We identified resected pathologic stage III NSCLC. For the radiation group (RT) group, at least 45 Gy of external beam radiation was required. mRNA was extracted from primary tumor. The predefined cut-point was the median RSI with a primary endpoint of local control. Similar criteria were then applied to two extramural datasets (E1; E2) with progression free survival as the primary endpoint.

**Results:** Median follow-up from diagnosis was 23.5 months (range: 4.8-169.6 months). RSI was associated with time to local failure in the RT group with a two-year rate of local control of 80% and 56% between RS and RR groups, respectively p=0.02. RSI was the only variable found to be significant on Cox local control analysis (HR 2.9; 95% CI: 1.2-8.2; p=0.02). There was no significance of RSI in predicting local control in patients not receiving RT, p=0.48. A cox regression model between receipt of radiotherapy and RSI combining E1 and E2 showed that the interaction term was significant for PFS (3.7; 95% CI 1.4-10; p=0.009). A summary measure combining E1 and E2 showed statistical significance for PFS between RR and RS patients treated with radiotherapy (HR 2.7l; 95% CI 1.3-5.6; p=0.007) but not in patients not treated with radiotherapy (HR 0.94; 95% CI 0.5-1.78; p=0.86).

**Conclusions:** RSI appears to be predictive for benefit from adjuvant radiation. Prospective validation is required.

## INTRODUCTION

Despite careful preoperative staging, up to one-third of patients undergoing curative resection for non-small cell lung cancer (NSCLC) may have unforeseen involved mediastinal lymph nodes discovered at the time of thoracotomy. (1) (2) This regional involvement is classified as pathologic stage III and has a high recurrence risk. (3) Addition of adjuvant chemotherapy provides a modest absolute improvement of 5.8% in disease-free survival (DFS) at 5 years, but addition of postoperative radiotherapy (PORT) remains controversial. (4) A meta-analysis of ten randomized trials has suggested an overall detrimental effect in survival with PORT, particularly in patients with *p*N0 or *p*N1 disease. (5) (6) However, these trials used older radiation techniques which may have resulted in non-cancer related deaths. (7) (8) Unplanned exploratory subgroup analysis of a single trial reported a survival benefit of PORT for *p*N2, but not *p*N1 disease.(9) Recently presented data from the LUNG Art trial, indicates that adjuvant PORT may improve PFS but did not meet OS endpoints(10). Many of the patients on this trial had pN2 disease found at the time of resection only (not on up front imaging). It therefore becomes critical to better delineate who may benefit from PORT.

Using intrinsic tumor radiosensitivity may be a better means to selectively use PORT. In fact, radiosensitivity varies significantly between NSCLCs. (11) For instance, elegant gene expression testing is gradually superceding anatomic features as a predictive tool for adjuvant decision-making in intermediate-stage breast cancers. (12) A similar personalized classifier could have broad impact in community practice for resected stage III NSCLC.

To address this problem, our group created the radiosensitivity index (RSI)(13-15). From an original pool of over 7,000 genes, ten hub genes were identified as central to survival in cancer cell lines after ionizing radiation. These 10 genes were trained into an informative mathematical model, and have since been observed to be predictive for post-RT recurrence, including breast, rectal, esophageal, head and neck, glioblastoma, uterine, pancreas cancer, and oligometastatic disease(13,16-26). In the current study, we tested if RSI could identify patients most likely to benefit from PORT with improved clinical responses, in three independent retrospective consecutive cohorts of resected pathologic stage III NSCLC.

## MATERIALS AND METHODS

### Design and Patient selection

Archived tumors resected between 2000 and 2010 were screened for eligibility using an institutional database (27). Inclusion criteria included: pathologically confirmed Stage IIIA or IIIB non-small cell lung cancer at time of definitive operation, grossly negative margins, and clinical follow-up information available. No cases of malignant pleural effusion were included. Either 3D conformal or intensity-modulated radiation was acceptable. Approval was received from the institutional review board. Clinical data was recorded using individual chart review by investigators blinded to the RSI variable. Race and smoking status were recorded from a pretreatment patient questionnaire. Local recurrence was defined as recurrence in the irradiated field. If no event occurred, then cases were censored at the date of last clinical evaluation.

### External validation

For the E1 dataset, data was obtained from the public Director’s Challenge Consortium for the Molecular Classification of Lung Adenocarcinoma (28). To ensure probesets matched, only U133A were used. No overlapping cases with other datasets were identified. Inclusion criteria included: stage IIIA or IIIB non-small cell lung cancer, grossly negative margins, and clinical follow-up information available. Cases without survival data were excluded. Information on type of cancer operation, type of adjuvant chemotherapy, and type of adjuvant radiation was not consistently available. For E2, dataset was obtained from the GSE8894 (29). Inclusion criteria: stage IIIA or IIIB non-small cell lung cancer.

### RNA preparation and gene expression profiling

This has been previously described(30). Briefly, tissues were stored at −140°C after being snap frozen in liquid nitrogen. Serial sectioning of each sample was used to histologically evaluate tumor and malignant cells content before RNA extraction. Gene expression values from Affymetrix U133A CEL files were normalized using the robust multiarray average (RMA) algorithm (31). Raw gene expression data for other datasets were obtained from NCICB (E1) and GEO (E2 – GSE8894).

The methodology for calculation of RSI has been previously published. (15) Probesets used for each gene were the same as in previous studies. (14) (32) (33) (34) Briefly, each of the 10 genes in the assay was ranked according to gene expression [from the highest (10) to the lowest expressed gene (1)]. RSI was determined using the previously published ranked based linear algorithm. (14) (15)

### Statistical Methods

Each dataset was analyzed independently. The 50th percentile for RSI was predefined as the cut-point to dichotomize into RSI_-good_ radiosensitive (RS) and RSI_-poor_ radioresistant (RR), as in previous studies(19,20,26). This RSI variable was compared with prognostic variables and local control in the primary dataset. In E1 and E2, local control information was not available and progression free survival (PFS) was the primary endpoint. PFS was defined as any lung cancer recurrence or death, from the date of surgical operation. To test differences between groups, the Kruskal-Wallis, Pearson’s chi-square and Fisher’s exact tests were used when appropriate. Association of RSI with outcome was tested by log-rank. Hazard ratios (HR) were derived using proportional hazards. Statistical analyses were performed using JMP 13 (SAS Institute Inc, Cary, NC, USA). All statistical tests were two-sided.

## RESULTS

### Demographics

Clinical characteristics are listed in **Table 1**. The population consisted of 100 stage III patients, 58 receiving PORT and 42 not receiving PORT. The population was primarily Caucasians with prior smoking exposure and adenocarcinoma histology. There were no substantive differences in available clinicopathologic factors between patients that did or did not receive RT and between RS and RR cohorts. These variables included age, gender, operation type, pathologic nodal status (pN), histology subtype, grade, receipt of adjuvant chemotherapy, and presence of angiolymphatic invasion.

**Table 1.** Patient and Tumor Characteristics of Primary Dataset.

| Patient Characteristics | RT | No-RT | p value | RSI-good (RS) | RSI-poor (RR) | p value |
| --- | --- | --- | --- | --- | --- | --- |
|  | n (%) | n (%) |  | n (%) | n (%) |  |
| **Total** | 58 (58%) | 42 (42%) |  | 50 (50%) | 50 (50%) |  |
| **Age, Median (range)** | 67 (36-80) | 68 (43-82) | 0.28 | 69 (36-82) | 66 (46-79) | 0.08 |
| **Race: Caucasian** | 56 (97%) | 41 (98%) | 0.68 | 50 (100%) | 47 (94%) | 0.21 |
| **Other<sup>‡</sup>** | 2 (3%) | 1 (2%) |  |  | 3 (6%) |  |
| **Gender: Female** | 24 (41%) | 25 (60%) | 0.07 | 24 (48%) | 25 (50%) | 0.84 |
| **Smoking: Current Smoker** | 17 (29%) | 10 (24%) | 0.57 | 12 (24%) | 15 (30%) | 0.45 |
| **Former Smoker** | 36 (62%) | 30 (71%) |  | 33 (66%) | 33 (66%) |  |
| **Never-Smoker** | 5 (9%) | 2 (5%) |  | 5 (10%) | 2 (4%) |  |
| **Histology of NSCLC** |  |  |  |  |  |  |
| **Adenocarcinoma** | 33 (57%) | 27 (64%) | 0.75 | 27 (54%) | 33 (66%) | 0.45 |
| **Squamous Cell** | 17 (29%) | 10 (24%) |  | 16 (32%) | 11 (22%) |  |
| **Other** | 8 (14%) | 5 (12%) |  | 7 (14%) | 6 (12%) |  |
| **Angiolymphatic Invasion Y** | 27 (47%) | 26 (62%) | 0.13 |  |  |  |
| **Tumor grade: 1 or 2** | 26 (45%) | 21 (50%) | 0.61 | 20 (40%) | 27 (54%) | 0.16 |
| **3** | 32 (55%) | 21 (50%) |  | 30 (60%) | 23 (46%) |  |
| **AJCC pN stage: 0** | 8 (14%) | 8 (19%) | 0.76 | 7 (14%) | 9 (18%) | 0.58 |
| **1** | 6 (10%) | 4 (10%) |  | 4 (8%) | 6 (12%) |  |
| **2** | 43 (72%) | 30 (71%) |  | 39 (78%) | 34 (68%) |  |
| **3** | 1 (2%) |  |  |  | 1 (2%) |  |
| **Surgery: Pneumonectomy** | 11 (19%) | 13 (31%) | 0.21 | 11 (22%) | 13 (26%) | 0.68 |
| **Lobectomy** | 35 (60%) | 23 (55%) |  | 30 (60%) | 28 (56%) |  |
| **Segmentectomy** | 4 (7%) |  |  | 1 (2%) | 3 (6%) |  |
| **Wedge Resection** | 8 (14%) | 6 (14%) |  | 8 (16%) | 6 (12%) |  |
| **Adjuvant Chemotherapy Y** | 56 (97%) | 42 (100%) | 0.14 | 49 (98%) | 49 (98%) | 1 |
| **Radiation Dose Median (Range) (Gy)** | 54.9 (43.2-70) |  |  | 54.9 (43.2-70) | 54.9 (45-70) | 0.7 |

\*RSI= Radiosensitivity index; \*\*RS= radiosensitive; \*\*\* RR= radioresistant; \*\*\*\*RT = radiotherapy

Median radiation dose to the planned target volume was 54.9 Gy (range: 43.2 – 70 Gy) in a median of 30 fractions (range: 23-36). No significant difference in median radiation dose was detected between the RS and RR groups (54.9 *vs*. 54.9 Gy, *p* = 0.7).

### Association with local control

The median follow-up from diagnosis was 23.5 months (range: 4.8-169.6 months). Median OS for the cohort was 42 months (95% CI 34-84 months) with a two-year OS rate of 75%. Median local control was 74 months (95% CI 31 months – not reached) with a 60% event-free rate at 5 yrs. RSI was associated with time to local failure in the RT group with a two-year rate of local control of 79% and 55% between RS and RR groups, respectively p=0.01, **Figure 1a**. RSI was the only variable found to be significant on Cox analysis (HR 2.9; 95% CI: 1.2-8.2; p=0.02); **Table 2**. There was no significance of RSI in predicting local control in patients not receiving RT with a two-year rate of local control of 68% and 73%, p=0.54, between RS and RR groups, respectively, **Figure 1b**.

**Table 2:** Univariable Model Cox Proportional Hazard Model.

| **Variable** | **Univariable Model** |  |
| --- | --- | --- |
|  | **HR (95% CI)** | **P-Value** |
| **RSI poor/good** | 2.9 (1.2-8.2) | 0.02 |
| **Tumor Grade 3/1-2** | 1.1 (0.5-2.5) | 0.8 |
| **pN 2/0-1** | 2.2 (0.7-9.2) | 0.17 |
| **Age ≥67/<67** | 0.52 (0.24-1.4) | 0.22 |

\*RSI= Radiosensitivity index

**Figure 1:**
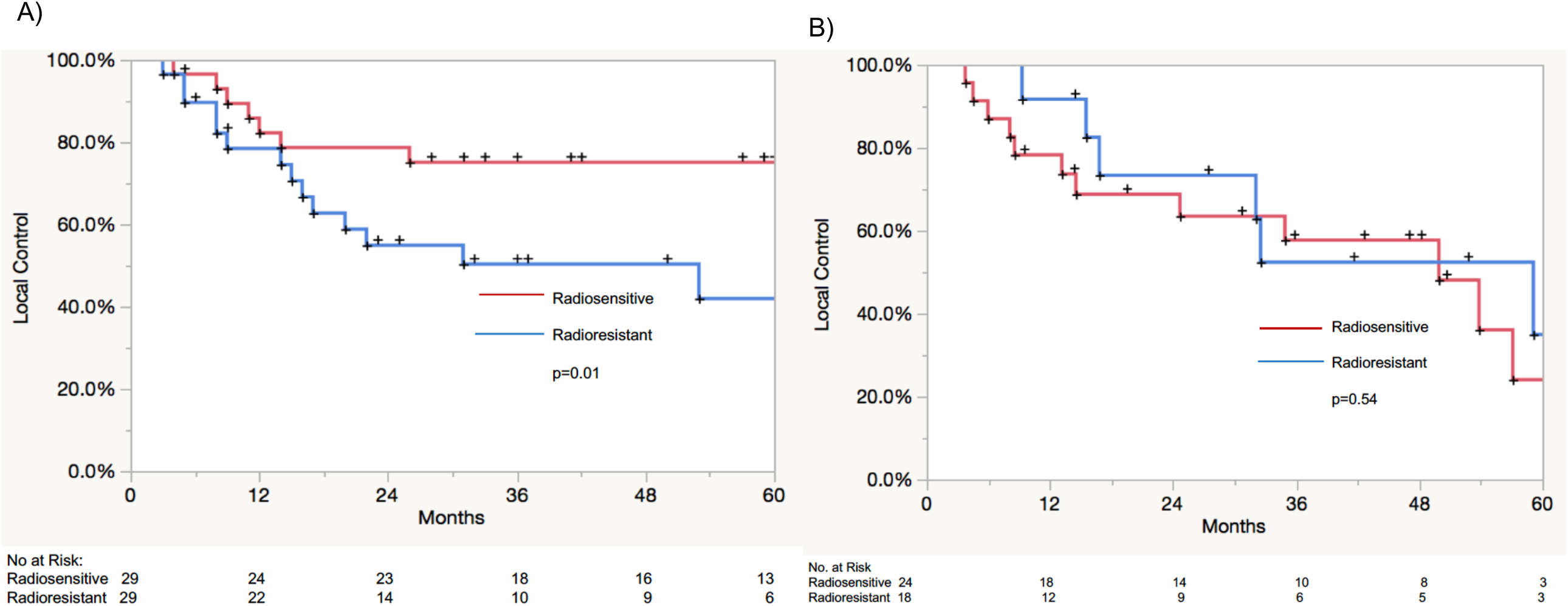
Local control between radiosensitive and radioresistant tumors A) receiving RT B) not receiving RT.

### External validation

Additional datasets with available gene expression data were queried after systematic review. Two sets were identified (E1, E2) and similar inclusion criteria applied. Both sets contained tumors archived when adjuvant chemotherapy was not in common practice.

For E1, subjects were primarily Caucasians with lung adenocarcinoma resected between 1992 and 2002 (**Table 3**). Differences were observed between RT and control for known covariates, including a higher proportion of RT patients receiving adjuvant chemotherapy (*p* <0.001), tumors that were grade 3 (p=0.04) and never smokers (p=0.02). No significant differences were noted between RS and RR cohorts. Median PFS in stage III patients was 13 months (95% CI 9-21 months) with a two-year PFS of 35% with a median follow-up of 22.1 months (range: 0.5-121.2 months). A trend towards inferior PFS for RR compared to RS cohorts receiving chemotherapy was observed in the RT group with a two-year rate of 10% and 50%, respectively (p=0.10). No trend was observed in the patients not receiving RT, RR and RS cohorts, 53% vs. 32% p=0.27.

**Table 3.** Patient and Tumor Characteristics of E1.

| Patient Characteristics | RT | No-RT | p value | RSI-good | RSI-poor | p value |
| --- | --- | --- | --- | --- | --- | --- |
|  | n (%) | n (%) |  | n (%) | n (%) |  |
| **Total** | 23 | 36 |  | 30 | 29 |  |
| **Age, median (range)** | 64 (48-78) | 64.5 (38-79) | 0.71 | 64.5 (48-79) | 64 (38-79) | 0.59 |
| **Race: Caucasian** | 21 (91%) | 24 (67%) | 0.2 | 23 (77%) | 22 (76%) | 0.53 |
| **Other<sup>‡</sup>** | 2 (9%) | 12 (33%) |  | 7 (23%) | 7 (24%) |  |
| **Gender: Female** | 12 (52%) | 15 (42%) | 0.43 | 17 (57%) | 10 (34%) | 0.09 |
| **Smoking: Current smoker** | 1 (4%) | 3 (8%) | 0.02 | 4 (13%) | 0 | 0.12 |
| **Former smoker** | 2 (9%) | 5 (14%) |  | 17 (57%) | 18 (62%) |  |
| **Never-smoker** | 19 (83%) | 16 (44%) |  | 3 (10%) | 4 (14%) |  |
| **Unknown** | 1 (4%) | 12 (33%) |  | 6 (20%) | 7 (24%) |  |
| **Histology of NSCLC** |  |  |  |  |  |  |
| **Adenocarcinoma** | 23 (100%) | 36 (100%) |  | 30 (100%) | 29 (100%) |  |
| **Tumor grade: 1 or 2** | 8 (36%) | 23 (64%) | 0.04 | 17 (57%) | 14 (50%) | 0.61 |
| **3** | 14 (64%) | 13 (36%) |  | 13 (43%) | 14 (50%) |  |
| **AJCC <sup>§</sup> pN stage:** |  |  |  |  |  |  |
| **1** | 1 (4%) | 7 (29%) | 0.1 | 5 (17%) | 3 (10%) | 0.48 |
| **2** | 22 (96%) | 29 (81%) |  | 25 (83%) | 26 (90%) |  |
| **Margin** |  |  |  |  |  |  |
| **R0** | 22 (96%) | 33 (92%) | 0.54 | 28 (93%) | 27 (93%) | 0.97 |
| **R1** | 1 (4%) | 3 (8%) |  | 2 (7%) | 2 (7%) |  |
| **Adjuvant Chemotherapy Y** | 18 (78%) | 3 (8%) | <0.001 | 10 (33%) | 11 (38%) | 0.71 |

\*RSI= Radiosensitivity index; \*\*RS= radiosensitive; \*\*\* RR= radioresistant; \*\*\*\*RT = radiotherapy

For E2, subjects were Asians with NSCLC resected between 1998 and 2005 (**Table 4)**. A significant difference was noted in the receipt of adjuvant chemotherapy between the RT and non-RT cohorts. No differences were noted in RR and RS cohorts. Median PFS was 5.2 months (95% CI: 3.8-11 months) with median follow-up of 21.7 months. Similar to E1, the RR subjects had a trend towards inferior PFS in the RT group compared to RS patients, with a 12 month PFS rate of 40% and 55%, p=0.14, respectively. In patients, not treated with RT there was no trend observed, p=0.85.

**Table 4.** Patient and Tumor Characteristics of E2.

| Patient Characteristics | RT | No-RT | p value | RSI-good | RSI-poor | p value |
| --- | --- | --- | --- | --- | --- | --- |
|  | n (%) | n (%) |  | n (%) | n (%) |  |
| **Total** | 16 (62%) | 10 (38%) |  | 13 (50%) | 13 (50%) |  |
| **Age, median (range)** | 58 (40-74) | 51.5 (13-75) | 0.37 | 57 (40-72) | 54 (13-75) | 0.86 |
| **Race: Asian** | 16 (100%) | 10 (100%) |  | 13 (100%) | 13 (100%) |  |
| **Gender: Female** | 3 (19%) | 0 | 0.15 | 1 (8%) | 2 (15%) | 0.54 |
| **Smoking** |  |  |  |  |  |  |
| **Current/Former smoker** | 11 (69%) | 9 (90%) | 0.19 | 11 (85%) | 9 (69%) | 0.35 |
| **Never-smoker** | 5 (31%) | 1 (10%) |  | 2 (15%) | 4 (31%) |  |
| **Surgery** |  |  |  |  |  |  |
| **Pneumonectomy** | 6 (38%) | 6 (60%) | 0.42 | 5 (38%) | 7 (54%) | 0.72 |
| **Lobectomy** | 9 (56%) | 3 (30%) |  | 7 (54%) | 5 (38%) |  |
| **Bilobectomy** | 1 (6%) | 1 (10%) |  | 1 (8%) | 1 (8%) |  |
| **Histology of NSCLC** |  |  |  |  |  |  |
| **Adenocarcinoma** | 8 (50%) | 4 (40%) | 0.62 | 6 (46%) | 6 (46%) | 1 |
| **Squamous Cell** | 8 (50%) | 6 (60%) |  | 7 (53%) | 7 (53%) |  |
| **Tumor grade: 1 or 2** | 9 (56%) | 8 (80%) | 0.2 | 7 (54%) | 10 (77%) | 0.27 |
| **3** | 7 (44%) | 2 (20%) |  | 6 (46%) | 3 (23%) |  |
| **AJCC <sup>\$</sup> pN stage:** | | | | | | |
| **0** | 1 (6%) | 4 (40%) | 0.09 | 1 (8%) | 4 (31%) | 0.31 |
| **1** | 7 (44%) | 2 (20%) |  | 5 (38%) | 4 (31%) |  |
| **2** | 8 (50%) | 4 (40%) |  | 7 (54%) | 5 (38%) |  |
| **Adjuvant Chemotherapy Y** | 0 | 4 (40%) | 0.006 | 1 (8%) | 3 (23%) | 0.27 |

\*RSI= Radiosensitivity index; \*\*RS= radiosensitive; \*\*\* RR= radioresistant; \*\*\*\*RT = radiotherapy

A Cox regression model between receipt of radiotherapy and RSI combining E1 and E2 showed that the interaction term was significant for PFS (3.7; 95% CI 1.4-10; p=0.009). A summary measure combining E1 and E2 showed statistical significance for PFS between RR and RS cohorts in patients treated with radiotherapy (HR 2.7l; 95% CI 1.3-5.6; p=0.007) but not in patients not treated with radiotherapy (HR 0.94; 95% CI 0.5-1.78; p=0.86), **Figure 2**.

**Figure 2:**
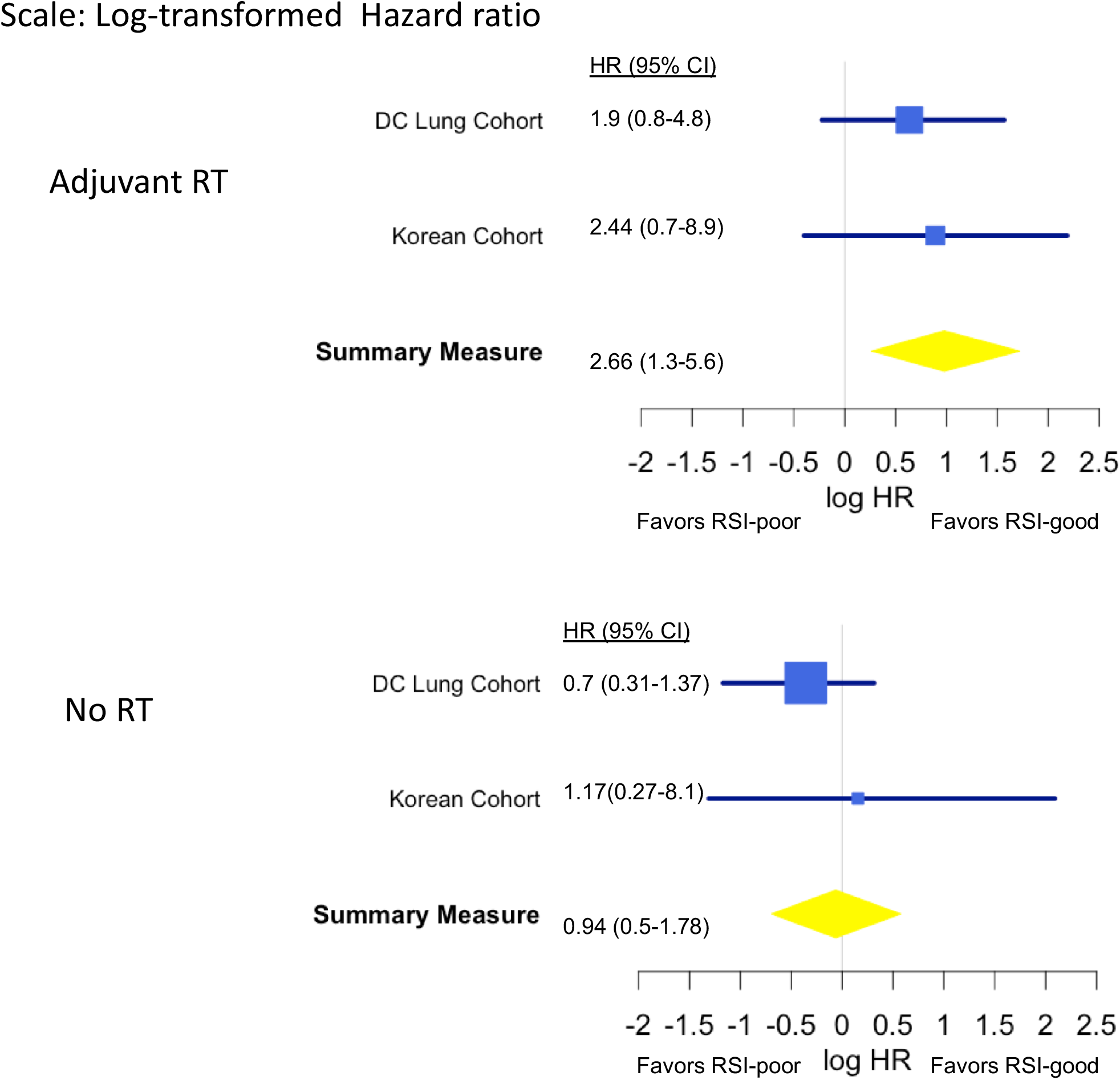
Forest plot of progression free survival between E1 and E2.

## DISCUSSION

The decision to give adjuvant PORT is controversial and balancing risks vs. toxicity is key. To date, no predictive gene-based tool has been implemented for PORT in NSCLC. In the current report, we tested a signature that has been previously authenticated in twelve previously published independent datasets with other cancers. Although a retrospective analysis, the study used a pre-specified gene signature with a predetermined cut-point, with the same normalization method. Robust covariates were collected, such as surgery type, grade and histology, to assess for potential confounding. We did not detect substantive imbalances in these prognostic factors, and RSI remained an important predictor even after accounting for these factors. Furthermore, the study tested two independent radiation datasets, albeit small in size, and the point estimate remained consistent across all three. Finally, the predictive significance of the assay was bolstered by the absence of any effect in the negative control groups.

Although prognostic gene-based signatures are abound for resected NSCLC, comparatively few predictive signatures for radiation benefit have been reported. (35) However, multiple genes in apoptosis and DNA repair have been proposed to mediate innate and acquired radioresistance. For example, up-regulation of MDM2 may correlate with radioresistance in resected squamous NSCLC, and antisense suppression of MDM2 may restore radiosensitivity in A549 cells. (36) (37) Along these lines, an inducible suppressor of p53, called tumor protein p53 inducible protein 3 (TP54I3), is linked with radiosensitivity in H460 and A549 cells. (38) Normal cancer cells had radiation–induced PARP cleavage, whereas radioresistant cells did not. Plasma detection of the DNA repair enzymes ERCC and XRCC was correlated with OS in RTOG 0617, a radiation dose-escalation trial in NSCLC. (39) Consistent with these reports, the 10 genes of RSI are also mediators of DNA repair, cell cycle, and apoptosis.

There is growing evidence to demonstrate the value of RSI in guiding radiotherapy decisions in a number of malignancies(16,19-22,40-42). RSI has been previously found to identify radioresistant triple negative breast cancer tumors at higher risk for local recurrence compared to radiosensitive triple negative tumors(22). Strom et al. demonstrated that pancreatic cancer patients with high risk features (positive post-operative margin, positive lymph nodes, or post-operative CA19-9 > 90) receiving RT could be stratified by RSI. In the study, RSI-poor (RR) tumors had significantly worse outcomes compared to RSI-good (RS) tumors with a median OS of 13.2 compared to 31.2 months, respectively (*p*=0.04).(20) The principle of these findings and those of the current study in post-operative delivery of lung radiotherapy were also demonstrated in studies regarding uterine carcinoma, lung metastases, colon primaries and metastases, and glioblastoma.(19,21,25,26,41)

This study is characterized by important limitations inherent to a retrospective report. Not all patients received anatomic lobar resections, a condition commonly encountered in practice. Moreover, some patients received pneumonectomy, which is a risk factor for poor outcome after PORT. (43) Since this was a study of resected NSCLC, response rate was not possible to use as an endpoint. Along these lines, radiation treatment plans were not available to review, in the external datasets and we were left with the endpoint of PFS for analysis while local control was used for the primary dataset. Similarly, the frequency of follow-up intervals and CT imaging were not absolutely uniform, since this was a non-randomized, retrospective study. (44) Specifically, most patients received CT scans every 3 to 4 months in their first two years after surgery. Furthermore, since radiation was not a randomized assignment, comparison tests for outcome of RT *vs*. control group were not possible. Thus, we did not report a theoretical risk reduction for RSI, as it might be used in a clinical trial. Another limitation is that relatively small sample sizes were accessible for the present study. It seems that locally advanced NSCLC may be underrepresented in available datasets, and annotation regarding adjuvant radiation is often omitted or unaccounted for.

RSI deserves additional investigation, ideally in a prospective manner. Given the controversy and adverse effects of adjuvant radiation, a tool to inform patient selection for adjuvant RT or dose could be informative for radiation oncology practice. In conclusion, using a previously validated genomically-derived radiation specific signature, we have identified a subset of lung cancer patients with RR tumors that are at higher risk for failure. We propose RSI may be a means to identify post-operative lung patients at higher risk for failure and more likely to derive benefit from PORT.

## Data Availability

All data can be made available upon request.

## Acknowledgements

Dr. Sumin Shin and Dr. Jhingook Kim from the Samsung Medical Center, Sungkyunkwan University School of Medicine provided data for the study.

